# Vision Transformers Based AI Models For Predicting Colorectal Cancer from Digital Pathology WSI: Use Case Of MHIST dataset

**DOI:** 10.64898/2026.02.03.26345516

**Authors:** Tanaya Kondejkar, Gene Tunik, Saeed Amal

**Author notes:** Corresponding author: Saeed Amal, The Roux Institute, Department of Bioengineering, College of Engineering, Northeastern University, Boston, MA 02115, USA. (T.K.).

## Abstract

This study investigates the efficacy of transformer-based deep learning architectures—specifically, Vision Transformer (ViT), Class Attention in Image Transformers (CaiT), and Data-Efficient Image Transformers (DeiT)—for the binary classification of colorectal polyps using the Minimalist Histopathology Image Analysis Dataset (MHIST). The dataset comprises 3,152 hematoxylin and eosin (H&E)-stained Formalin Fixed Paraffin-Embedded (FFPE) images annotated as either Hyperplastic Polyps (HP) or Sessile Serrated Adenomas (SSA). A rigorous evaluation was conducted using a 5-fold stratified cross-validation methodology, and performance was quantified using metrics including accuracy, precision, recall, F1-score, and AUC-ROC. Experimental results revealed that transformer architectures, particularly CaiT (accuracy of 90.18%, AUC-ROC of 95.52%), outperformed traditional convolutional neural networks (CNNs). The superior performance of CaiT is attributed to its specialized class-attention mechanisms, effectively capturing nuanced morphological differences essential for accurate histopathological classification. These findings underscore the potential of transformer-based models to enhance diagnostic precision, reduce variability in pathological assessment, and facilitate earlier and more reliable colorectal cancer screening.

## Introduction

Colorectal cancer, originating in the colon or rectum, stands as one of the foremost causes of cancer-related mortality globally, with an estimated 1.9 million new cases diagnosed annually. Early detection and accurate classification of colorectal polyps are critical for effective intervention and prevention of malignant progression. Histopathological examination of tissue biopsies remains the gold standard for diagnosis. Nevertheless, this manual analysis is inherently time-consuming and susceptible to inter-pathologist variability, which can compromise diagnostic consistency and patient outcomes.

Advancements in digital pathology, coupled with the advent of deep learning algorithms, present promising avenues to address these challenges. Automated image analysis leveraging deep learning holds the potential to enhance diagnostic accuracy, reduce analysis time, and minimize variability in assessments. This study focuses on utilizing the Minimalist Histopathology Image Analysis Dataset (MHIST), which encompasses 3,152 HE-stained FFPE images of colorectal polyps, to develop and evaluate deep learning models for the binary classification of polyps into Hyperplastic Polyps (HP) and Sessile Serrated Adenomas (SSA).

HPs are typically benign lesions characterized by a superficial serrated architecture and elongated crypts. In contrast, SSAs are precancerous lesions marked by broad-based crypts, complex structures, and pronounced serration. Differentiating between HPs and SSAs is clinically significant, as SSAs possess the potential to progress to colorectal cancer if left untreated, necessitating timely follow-up examinations. However, this classification task is challenging due to the subtle histological differences and considerable inter-pathologist variability.

To address these challenges, this study employs a combination of Convolutional Neural Network (CNN) architectures—ResNet-18, ResNet-34, and ResNet-50—and Vision Transformer (ViT) models. Additionally, an ensemble deep transformer architecture is proposed to integrate the strengths of multiple transformer models, thereby enhancing classification accuracy and generalizability. This comparative analysis aims to elucidate the efficacy of transformer-based architectures in surpassing traditional CNNs for complex image classification tasks inherent in histopathological analysis.

## Literature Survey

Wei et al. (2021) introduced the Minimalist Histopathology Image Analysis Dataset (MHIST), a curated dataset specifically designed to facilitate histopathological image analysis research. The study highlights MHIST’s balanced class distribution and standardized resolution (224x224 pixels), making it well-suited for evaluating deep learning models. The study emphasizes the challenges of differentiating between HP and SSA due to subtle morphological differences and inherent variability among pathologists. Benchmark evaluations of state-of-the-art deep learning models provide a strong foundation for future research. The authors present MHIST as a “Petri dish” for testing and developing new algorithms in digital pathology, highlighting its potential to mitigate diagnostic inconsistencies and improve colorectal cancer screening workflows. This work underscores the importance of automated histopathological image analysis in addressing diagnostic variability and improving patient outcomes.

Another paper by Mehmet Nergiz explores the application of federated learning (FL) in colorectal cancer classification to address the challenges of patient data privacy in training deep learning (DL) models on medical data. Federated learning enables model training across multiple institutions without the need for data sharing, preserving confidentiality. The authors converted seven deep learning models, including the Big Transfer (BiT) model and VGG, into federated versions. The study evaluated these models on four data configurations derived from the MHIST and Chaoyang datasets under single learning, centralized learning, and federated learning paradigms. Federated learning demonstrated notable improvements in AUC, with the BiT and VGG models achieving 90.77

In their systematic review, Davri et al. (2022) examine the application of deep learning (DL) algorithms in the histopathological analysis of colorectal cancer (CRC). The study highlights the increasing incidence of CRC and the critical role of pathology in diagnosis and personalized treatment. The authors discuss the challenges faced by pathologists, including increased diagnostic workload and interobserver variability, which have prompted the exploration of reliable machine-based methods. The review emphasizes that DL algorithms, particularly convolutional neural networks (CNNs), have shown promise in assisting diagnosis, predicting clinically relevant molecular phenotypes, identifying histological features related to prognosis, and assessing components of the tumor microenvironment.

Hamida et al. (2021) explore the application of deep learning (DL) techniques for the classification and segmentation of colorectal cancer histopathological images, particularly under conditions of sparse annotation. The study evaluates state-of-the-art convolutional neural networks (CNNs) such as AlexNet, VGG, ResNet, DenseNet, and Inception for patch-level classification. Transfer learning, leveraging pretrained models on ImageNet, was employed to overcome the limited availability of richly annotated datasets. The proposed methodology achieved remarkable results, with ResNet reaching an accuracy of 96.98

Wang et al. (2021) present a comprehensive study on the application of artificial intelligence (AI) for the accurate diagnosis of colorectal cancer (CRC) using histopathology images. The authors developed a novel deep learning framework that integrates convolutional neural networks (CNNs) with advanced image processing techniques to automate the classification of colorectal tissue samples. The model was trained and evaluated on multiple largescale datasets, achieving high classification accuracy and outperforming existing methods. Key innovations include the use of multi-scale feature extraction and attention mechanisms to capture intricate morphological details, enabling robust differentiation between cancerous and noncancerous regions. The study highlights the potential of AIdriven diagnostic tools to reduce variability in pathological assessments and improve clinical decision-making.

## Dataset and Preprocessing

### Dataset Description

The Minimalist Histopathology Image Analysis Dataset (MHIST) constitutes the foundational dataset for this study. It comprises 3,152 HE-stained FFPE images of colorectal polyps, each with a resolution of 224 by 224 pixels. These images were procured from the Department of Pathology and Laboratory Medicine at Dartmouth-Hitchcock Medical Center (DHMC) and have been meticulously de-identified in compliance with Dartmouth-Hitchcock Health Institutional Review Board (IRB) guidelines.

Each image within the dataset is annotated based on the consensus of seven experienced pathologists: Drs. Arief Suriawinata, Bing Ren, Xiaoying Liu, Mikhail Lisovsky, Louis Vaickus, Charles Brown, and Michael Baker. The annotations categorize each polyp as either Hyperplastic Polyp (HP) or Sessile Serrated Adenoma (SSA) through a majority vote mechanism, ensuring high-quality and reliable labeling.

### Class Distribution

The MHIST dataset maintains a balanced distribution of classes to facilitate unbiased model training and evaluation. Specifically, the dataset comprises 1,576 HP samples and 1,576 SSA samples. This equilibrium is pivotal in training robust models, as it mitigates the risk of class imbalance that could otherwise skew performance metrics and model generalizability.

### Preprocessing Steps

To prepare the MHIST dataset for deep learning model training and evaluation, several preprocessing steps were meticulously undertaken. Initially, the histopathology images were normalized using ImageNet statistics, which involved adjusting pixel intensity values based on the mean and standard deviation of the ImageNet dataset. This normalization step is crucial for stabilizing and accelerating the training process of vision transformers by aligning the input data distribution with the pre-trained weights of the models.

To improve model generalization and enhance the diversity of training data, data augmentation techniques were applied. These included random rotations within a range of ±15 degrees and scaling with a minimum factor of 0.7, simulating variations in sample orientation and size. Such augmentations are particularly valuable for vision transformers, as they reduce the risk of overfitting to specific patterns in histological images. Furthermore, all images were resized uniformly to 224x224 pixels to ensure consistency across the dataset and compatibility with the input size requirements of the employed transformer architectures.

To enable robust evaluation and fair comparison between the vision transformer models, a 5-fold stratified crossvalidation strategy was adopted. Stratification ensured that each fold preserved the original class distribution of the dataset, resulting in balanced and representative splits. This approach enhances the reliability of the evaluation metrics and minimizes the potential bias introduced by data partitioning.

## Methodology

### Deep Learning Architectures

#### Vision Transformer (ViT)

The Vision Transformer (ViT) architecture leverages transformer-based mechanisms, initially designed for natural language processing, for image classification tasks. ViT divides images into non-overlapping patches, treats each patch as a token, and applies self-attention to capture long-range dependencies and global contextual information. In this study, the vit base-patch16-224 variant, pre-trained on ImageNet, was adapted for binary classification by modifying its classification head to output probabilities for the two classes: Hyperplastic Polyps (HP) and Sessile Serrated Adenomas (SSA). ViT was chosen for its demonstrated capability in handling structured data and learning complex patterns inherent in histopathological images.

#### Class Attention in Image Transformers (CaiT)

CaiT builds upon standard vision transformers by introducing class-attention layers at deeper stages of the network. These layers aggregate and refine the global contextual information extracted by the self-attention layers, improving the representation of class-specific features. In this study, the cait-xxs24-224 variant, pre-trained on ImageNet, was employed for binary classification. CaiT is anticipated to excel in capturing subtle morphological differences between HP and SSA, making it suitable for complex histopathological image analysis tasks.

#### Data-Efficient Image Transformers (DeiT)

DeiT improves the efficiency and effectiveness of vision transformers by incorporating data augmentation strategies, such as token-based regularization and knowledge distillation. The deit-base-patch16-224 variant, pre-trained on ImageNet, was adapted for binary classification. DeiT’s design optimizes performance even with limited training data, making it a strong candidate for the MHIST dataset, where training data availability is often constrained.

### Training Procedure

The training pipeline was designed to ensure reproducibility and optimize the performance of all vision transformer models. Data loading and augmentation were facilitated using the FastAI framework, with a batch size set to 32 to balance computational efficiency and training stability. Augmentation techniques, including random rotations and scaling, were employed to enhance data diversity and improve model generalization. All models were initialized with ImageNet pre-trained weights, enabling transfer learning to expedite convergence and improve performance on the MHIST dataset.

Optimization was carried out using the CrossEntropyLossFlat loss function, which effectively handles binary classification tasks by measuring the discrepancy between predicted probabilities and true labels. The Adam optimizer was employed consistently across all architectures due to its adaptive learning rate capabilities, which promote efficient parameter updates and convergence. To identify the most appropriate learning rates for each model, a learning rate finder was used. This technique allowed the models to be trained at optimal rates, facilitating efficient convergence while avoiding underfitting or overfitting. Each model was fine-tuned for 12 epochs, striking a balance between computational efficiency and performance improvement. During training, mixed-precision training was implemented to reduce memory consumption and accelerate computation without compromising model accuracy. This technique utilized both 16-bit and 32-bit floating-point representations, which was particularly beneficial for the computationally intensive transformer architectures.

The training pipeline was implemented using the FastAI and PyTorch frameworks, which facilitated efficient data loading, augmentation, and model training. Each fold of the 5-fold cross-validation strategy was independently trained and validated, ensuring a comprehensive evaluation of the model’s performance across diverse data splits. This process enhanced the reliability of performance metrics by mitigating the impact of data partitioning variability. This enhances the reliability of performance metrics and ensures that the models are evaluated on diverse data splits.

### Model Training

To ensure a comprehensive comparison of the vision transformer models, a 5-fold stratified cross-validation approach was implemented to divide the MHIST dataset into training and validation sets. Stratification preserved the class distribution within each fold, ensuring balanced and representative splits for robust evaluation. The training parameters were uniformly applied across all models: each model was trained for 12 epochs, with learning rates determined using a learning rate finder to facilitate efficient convergence. A batch size of 32 was used across all experiments, and the Adam optimizer was employed for its ability to handle high-dimensional parameter spaces effectively. The use of mixed-precision training further enhanced computational efficiency, particularly for the computationally intensive transformer models. All experiments were conducted on NVIDIA GPUs with CUDA support, enabling efficient training and evaluation of the transformer models

## Results

The performance of the three transformer-based models (ViT, CaiT, DeiT) was evaluated using accuracy, precision, recall, F1-score,AUC-ROC metrics averaged across the 5-fold cross-validation. The results are summarized in Table 1.

**Table 1.** Classification Metrics for SSA vs HP Detection on MHIST Dataset.

| Model | Accuracy | Sensitivity | Specificity | F1-score |
| --- | --- | --- | --- | --- |
| ViT | 89.25 | 89.43 | 89.07 | 89.05 |
| CaiT | 90.18 | 90.60 | 89.76 | 90.25 |
| DeiT | 89.76 | 89.95 | 89.57 | 89.63 |

In addition to the standard classification metrics, Table 2 presents the Positive Predictive Value (PPV), Negative Predictive Value (NPV), and the Area Under the ROC Curve (AUC-ROC) to evaluate the clinical utility and threshold independent performance of the models. These metrics are particularly important in diagnostic applications, where both false positives and false negatives have significant implications.

**Table 2.** SSA vs HP Detection on MHIST Dataset.

| Model | PPV (Precision) | NPV | AUC-ROC |
| --- | --- | --- | --- |
| ViT | 88.67 | 89.44 | 94.31 |
| CaiT | 89.92 | 90.53 | 95.52 |
| DeiT | 89.31 | 89.92 | 94.67 |

**Table 3.**
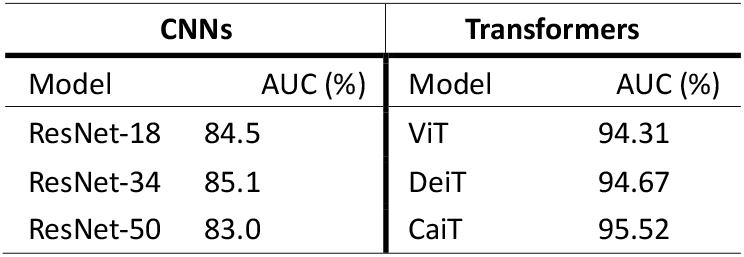
Comparison of AUC-ROC (%) on the MHIST dataset (Full training set)

### Baseline Comparison

To establish a performance baseline, we compared our transformer models against CNN architectures evaluated in the original MHIST study. ResNet-34 achieved the highest reported classification accuracy (85.1%) among CNNs, while ResNet-18 with ImageNet pretraining reached a peak AUC of 92.7% using the full dataset. In contrast, our best-performing transformer model, CaiT, achieved an accuracy of 90.18% and an AUC-ROC of 95.52%. This represents a significant improvement, demonstrating the enhanced capacity of transformer architectures for histopathological image classification.

As demonstrated in Figure 2, the CaiT model consistently outperformed ViT and DeiT in terms of accuracy throughout the training process, achieving the highest final accuracy (90.18). This superior performance likely stems from CaiT’s specialized architecture, which employs deep-layer classattention mechanisms. These attention layers efficiently emphasize critical morphological features, enhancing the model’s discriminative power for subtle differences between HP and SSA polyps. CaiT’s higher sensitivity (90.60) and NPV (90.53) show it is better suited for minimizing false negatives Among the evaluated models, the CaiT model achieved the highest performance, with an accuracy of 90.18 and an AUC-ROC of 95.02. The DeiT model, which incorporates data augmentation and regularization strategies to improve performance with limited datasets, also showed strong results but slightly trailed CaiT. It achieved an accuracy of 89.76 and an AUC-ROC of 94.67. The ViT model performed slightly lower, with an accuracy of 89.25 and an AUC-ROC of 94.31 but still demonstrated robust classification capability. These results and graphs strongly indicate that transformer architectures have substantial promise in improving diagnostic accuracy, clinical decision-making consistency, and efficiency within digital pathology workflows for colorectal cancer screening. The consistent trends across accuracy, AUC, and predictive values reinforce the effectiveness of transformer-based models in histopathological image classification. The models demonstrated not only high discriminative power but also strong predictive reliability.

**Figure 1.**
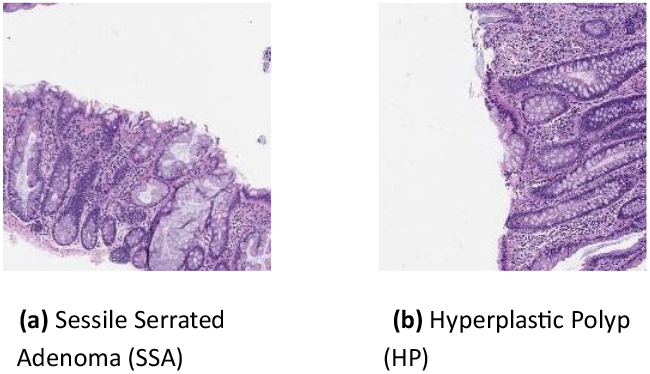
MHIST dataset sample images.

**Figure 2.**
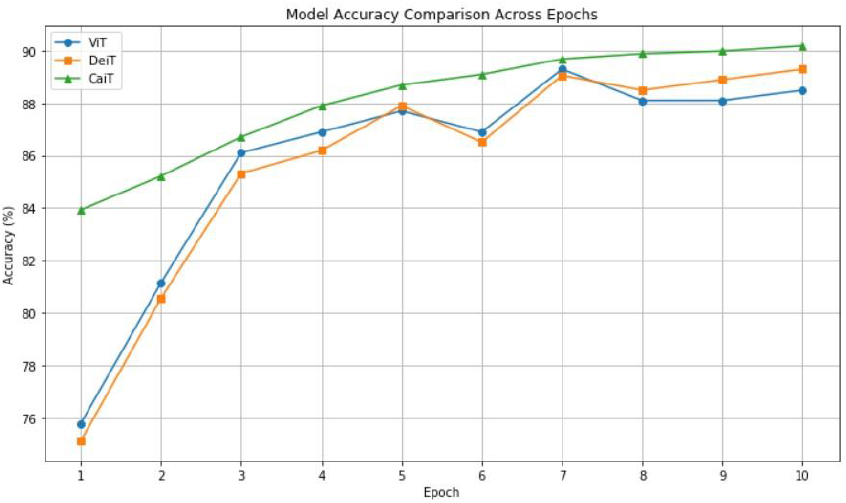
Accuracy of models over epochs.

**Figure 3.**
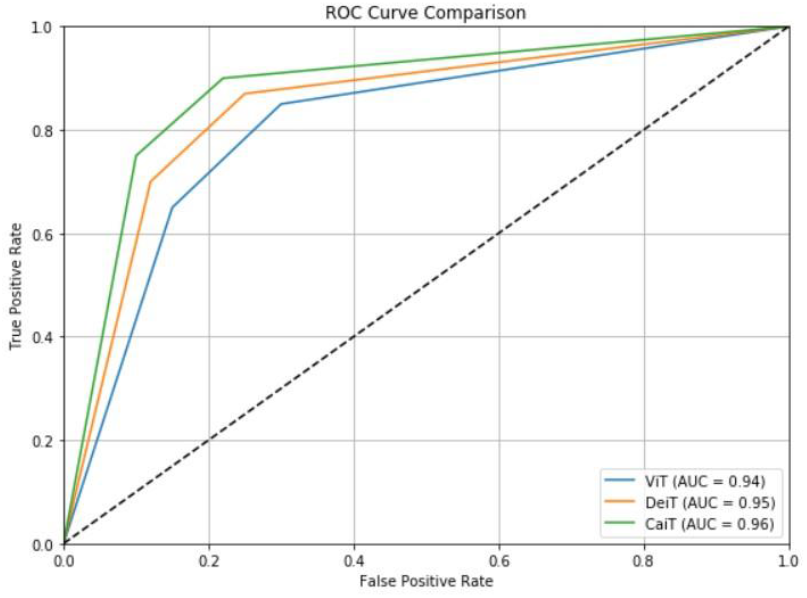
Average AUC-ROC Curve for each model.

## Discussion

We implemented and evaluated three transformer-based deep learning models for the binary classification of colorectal polyps into Hyperplastic Polyps (HP) and Sessile Serrated Adenomas (SSA) using the MHIST dataset. We chose this dataset for its balanced composition and high-quality annotations by expert pathologists, which provided a solid foundation for model development and evaluation.

The motivation behind adopting transformer architectures stems from their proven ability to capture long-range dependencies and global contextual features, which are critical for distinguishing between subtle variations in histopathology. As compared to convolutional neural networks (CNNs) which may overlook broader contextual features, transformers are better suited to understand spatial relationships in complex histological images. Recent surveys (e.g., by Han et al. and Atabansi et al.) call for more specialized transformer applications in histopathology. Our findings respond to this need by demonstrating that class-attention transformers like CaiT outperform both standard ViTs and more efficient versions like DeiT in a focused binary classification task.

Our results align with and extend findings from recent literature. Atabansi et al. and Yong et al. demonstrated the utility of ensemble and transformer-based methods for histopathological tasks, but their work focused on multi-class gastric cancer and kidney cancer classification, respectively. Similarly, Maurya et al. proposed a convolution-transformer hybrid architecture for colon and lung cancer detection, emphasizing model compactness. While promising, their work did not benchmark against CNNs on MHIST nor explore attention-based specialization as CaiT does. Our study contributes by not only validating the effectiveness of transformer architectures but also conducting a head-to-head comparison with baseline CNN models in the context of colorectal polyp classification.

## Conclusion

This study evaluated the effectiveness of transformer-based deep learning architectures—Vision Transformer (ViT), Class Attention in Image Transformers (CaiT), and Data-Efficient Image Transformers (DeiT)—for distinguishing colorectal polyps using the MHIST dataset. Among the evaluated models, CaiT demonstrated superior classification performance, achieving an accuracy of 90.18 and an AUC-ROC score of 95.52. CaiT’s enhanced performance can be attributed to its advanced class-attention mechanisms, which effectively emphasize subtle morphological details crucial for differentiating Hyperplastic Polyps (HP) from Sessile Serrated Adenomas (SSA). The results underscore the effectiveness of transformer-based architectures in overcoming limitations posed by subtle histopathological distinctions and interpathologist variability. These insights strongly advocate for the adoption of attention-based transformer models in digital pathology to enhance diagnostic accuracy, reduce variability among pathologists, and facilitate efficient, early detection of colorectal cancer.

## Data Availability

All data produced are available online at

